# Comparative Performance of SARS-CoV-2 Detection Assays using Seven Different Primer/Probe Sets and One Assay Kit

**DOI:** 10.1101/2020.03.13.20035618

**Authors:** Amanda M. Casto, Meei-Li Huang, Arun Nalla, Garrett Perchetti, Reigran Sampoleo, Lasata Shrestha, Yulun Wei, Haiying Zhu, Alexander L. Greninger, Keith R. Jerome

## Abstract

More than 100,000 people worldwide are known to have been infected with SARS-CoV-2 beginning in December 2019. The virus has now spread to over 93 countries including the United States, with the largest cluster of US cases to date in the Seattle metropolitan area in Washington. Given the rapid increase in the number of local cases, the availability of accurate, high-throughput SARS-CoV-2 testing is vital to efforts to manage the current public health crisis. In the course of optimizing SARS-CoV-2 testing performed by the University of Washington Clinical Virology Lab (UW Virology Lab), we tested assays using seven different primer/probe sets and one assay kit. We found that the most sensitive assays were those the used the E-gene primer/probe set described by Corman et al. (Eurosurveillance 25(3), 2020, https://doi.org/10.2807/1560-7917.ES.2020.25.3.2000045) and the N2 set described by the CDC (Division of Viral Diseases, Centers for Disease Control and Prevention, 2020, https://www.cdc.gov/coronavirus/2019-ncov/downloads/rt-pcr-panel-primer-probes.pdf). All assays tested were found to be highly specific for SARS-CoV-2, with no cross-reactivity with other respiratory viruses observed in our analyses regardless of the primer/probe set or kit used. These results will provide invaluable information to other clinical laboratories who are actively developing SARS-CoV-2 testing protocols at a time when increased testing capacity is urgently needed worldwide.

## Background

In late December 2019, a cluster of cases of pneumonia of unclear etiology was first noted in Wuhan City in the Hubei Province of China (1). The etiology of these pneumonia cases, a novel type of coronavirus, was identified on January 7, 2020 (1). This novel coronavirus has now been named severe acute respiratory syndrome coronavirus 2 (SARS-CoV-2) while the disease it causes is known as coronavirus disease 2019 (COVID-19) (2).

To date, the epidemic has been largely concentrated in China, with a total of 80,695 known cases as of March 7, 2020 (3). However, cases outside of China were observed early in the epidemic with the first detected in Thailand on January 13 (4). Soon afterwards cases were also identified in other east Asian countries including Japan and South Korea (1), which now has the largest number of known cases outside of China with 7,134 as of March 7 (3).

The Centers for Disease Control and Prevention (CDC) confirmed the first case of COVID-19 in the United States on January 21. The infected person was a 35 year old man who had recently returned to his home in Snohomish County, Washington after travelling to Wuhan City (5). No additional cases of COVID-19 were identified in Washington State until February 28 when two new cases were confirmed, one in Snohomish County and one in neighboring King County, where Seattle is located (6). Since February 28, the number of cases of COVID-19 in Washington has steadily increased and currently stands at 102 (7).

In response to the rapidly increasing number of confirmed and suspected cases of COVID-19 in the Seattle metropolitan area, the Clinical Virology Laboratory at the University of Washington (UW Virology Lab) has begun testing clinical specimens for SARS-CoV-2. Prior to and since making this testing service available, we have endeavored both to optimize the performance of our assay and to increase the rate at which we are able to test samples. We report here our observations comparing three different RNA extraction methods. We also compare the performance of SARS-CoV-2 detection assays using seven different primer/probe sets and one assay kit.

## Materials and Methods

### Samples

Three sets of samples were used in our analyses. First, we used a set of approximately 300 clinical respiratory samples sent to the UW Virology Lab for respiratory virus testing. These samples were in the form of nasopharyngeal or oropharyngeal swabs in viral transport media and had not previously been tested for SARS-CoV-2. Secondly, we used a collection of nasal swabs in viral transport media that are used to validate all assays performed at the UW Virology Lab. This collection includes samples positive for: rhinovirus (3 samples within the set), influenza B (2), influenza A (2), parainfluenza virus 1 (1), parainfluenza virus 3 (2), parainfluenza virus 4 (1), adenovirus (2), metapneumovirus (1), bocavirus (2), respiratory syncytial virus (2), coronavirus (25). The coronaviruses included in the sample set are non-SARS-CoV-2 samples. Twenty-two negative samples are also included in this sample set. Finally, we obtained a set of 10 samples confirmed to be positive for SARS-CoV-2 by the Washington State Department of Health (WSDOH) Public Health Laboratories. These samples were also all nasopharyngeal or oropharyngeal swabs immersed in viral transport media.

### RNA Extraction

RNA extraction from samples was performed using two different systems, the MagNA Pure LC 2.0 and the MagNA Pure 96 (Roche Lifesciences). For both systems, RNA extraction was performed according to the manufacturer’s instructions. For the MagNA Pure LC 2.0, 200 µL of each sample was subjected to extraction with an elution volume of 200 µL. For the MagNA Pure 96, 200 µL of each sample was subjected to extraction with an elution volume of either 50 or 100 µL. 5 µL of RNA in elution buffer was used in each SARS-CoV-2 detection assay.

### SARS-CoV-2 Detection Assays

We used a total of 7 different primer/probe sets in our SARS-CoV-2 detection assays. The University of Washington (UW) RdRp primer/probe set was designed by the UW Virology Lab. Three additional primer/probe sets were designed as described in Corman et al. (8); these will be referred to as the Corman N-gene, RdRp, and E-gene primer/probe sets. The Centers for Disease Control (CDC) N1, N2, and N3 sets were developed by the CDC and have been published on the CDC website (9).

For all of the above primer/probe sets, real-time RT-PCR assays were performed using the AgPath-ID One Step RT-PCR kit (Life Technologies). 25 µL of reaction mix consists of 2x RT-PCR buffer, 25x enzyme mix, primers/probes, and 5 µL of extracted nucleic acid. Primer/probe concentrations were as recommended in Corman et al (8) and by a CDC recommended protocol (10). RT-PCR was performed on an ABI 7500 real-time PCR system (Applied Biosystems) with cycle parameters: 10 minutes at 48 °C for reverse transcription, 10 minutes inactivation at 95 °C followed by 40 cycles of 15 seconds at 95 °C and 45 seconds at 60 °C.

We also tested samples for SARS-CoV-2 using the BGI RT-PCR detection kit (BGI). These assays were conducted according to the kit manufacturer’s instructions.

A negative (human specimen control) was included in every RNA extraction procedure and non-template (water) control was included in every RT-PCR run. An internal control amplification was performed to monitor the extraction and RT-PCR quality.

## Results

### Assays using UW RdRp and Corman N-gene primer/probe sets found to have LODs of 1:2×10^3^

The first two primer/probes sets that we evaluated were the UW RdRp and the Corman N-gene sets. We tested the sensitivity of assays using these primer/probe sets by examining approximately 300 clinical samples of unknown SARS-CoV-2 status. RNA was extracted for these assays using two different systems. The first was the MagNA Pure LC (LC) system, which is able to process 32 samples at a time and elutes RNA into 200 µL of buffer. The second was the MagNA Pure 96 (MP96) system, which is able to process 96 samples at a time and elutes RNA into 100 µL of buffer. One sample out of the 300 was positive for SARS-CoV-2. The positive result was consistent regardless of which of the two RNA extraction methods and which of the two primer/probe sets was used.

We then determined the limit of detection (LOD) of assays using the UW RdRp and Corman N-gene primer/probe sets when run on dilutions of the one positive clinical sample. For both RNA extraction techniques and both primer/probe sets, the LOD was found to be 1:2×10^3^ (for each combination of RNA extraction method and primer/probe set, twenty out of twenty replicate assays were positive for SARS-CoV-2 when the positive clinical sample was diluted to 1:2×10^3^). Finally, we tested the specificity of assays using both primer/probe sets by running them on a collection of samples that are positive for respiratory viruses other than SARS-CoV-2. Assays using both sets were found to be 100% specific with no false positives noted in this analysis.

### Assays using the Corman RdRp and E-gene sets found to have LODs of 1:2×10^4^

We next tested assays using the Corman RdRp and Corman E-gene primer/probe sets. We again used two RNA extraction methods. The LC extraction method was performed using the same protocol as before. However, for the MP96 method, we eluted RNA into 50 µL of buffer instead of 100 µL of buffer to determine whether this would increase the sensitivity of SARS-CoV-2 detection assays. We ran assays using both the Corman RdRp and the Corman E-gene primer/probe sets coupled with both RNA extraction methods on 10 samples confirmed by the WSDOH Public Health Laboratories to be positive for SARS-CoV-2. The results of these tests are shown in Table 1. There was one sample that was positive for assays using the E-gene set when the MP96 system was used but not when the LC system was used and two samples that were positive for assays using the RdRp set when the MP96 system was used but not when the LC system was used. Based on these results, we subsequently used the MP96 RNA extraction system with RNA eluted into 50 µL of buffer for all analyses.

**Table 1:**
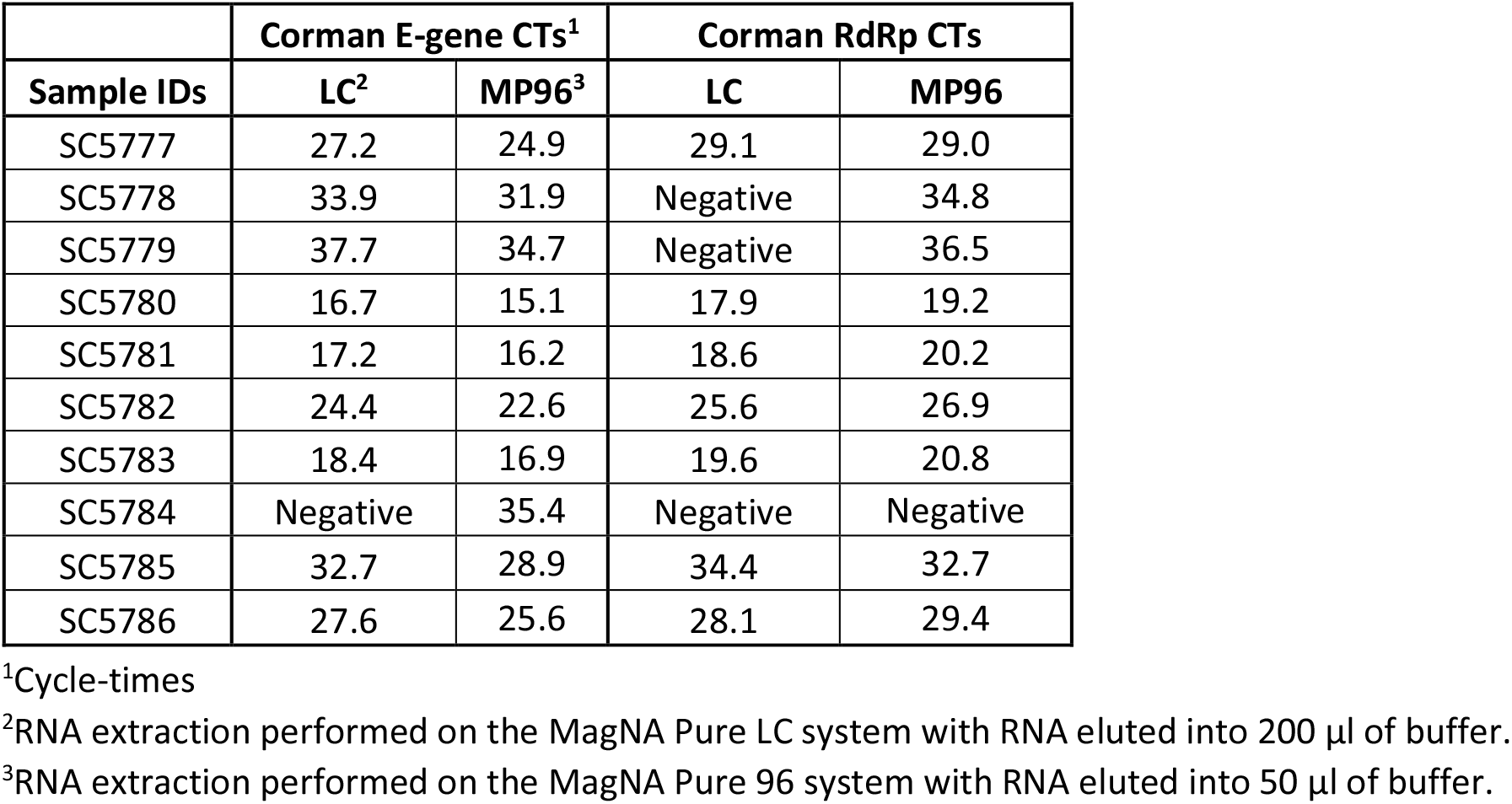
Relative performance of SARS-CoV-2 detection assays using the Corman E-gene and RdRp primer/probe sets and two different RNA extraction methods.

To assess the sensitivity of assays using the Corman RdRp and E-gene primer/probe sets, we ran them on dilutions of the positive UW Virology clinical sample. The LOD was found to be 1:2×10^4^ for both (again 20 replicate assays for each primer/probe set were performed and all 20 were positive). We also tested the specificities of assays using these sets by running them on our collection of samples positive for various respiratory viruses. Like assays using the UW RdRp and the Corman N-gene primers, those using the Corman RdRp and E-gene sets were 100% specific with no false positives noted.

### Assays using the CDC N1 and N2 primer/probe sets performed better those using the N3 set

Given that assays using the Corman RdRp and E-gene primer/probe sets showed superior performance relative to those using the UW RdRp and Corman N-gene sets, we wanted to compare the former to assays using the primer/probe sets published by the CDC: CDC N1, CDC N2, and CDC N3. We ran assays using these three sets on the 10 positive samples obtained from the WSDOH Public Health Laboratories. The results of these analyses are shown in Table 2. All assays produced positive results for all samples, except for the assay using the CDC N3 primers/probe which produced a negative result for SC5784. The assay using the Corman RdRp set also produced a negative result for this sample. For the other nine samples, assays using the Corman RdRp set consistently produced the highest cycle times out of all the assays compared in Table 2 (CDC N1, CDC N2, CDC N3, Corman RdRp, and Corman E-gene) followed by assays using the Corman E-gene set.

**Table 2:** Relative performance of SARS-CoV-2 detection assays using five different primer/probe sets.

| Sample IDs | CDC N1 | CDC N2 | CDC N3 | Corman RdRp | Corman E-gene |
| --- | --- | --- | --- | --- | --- |
| SC5777 | 24.5 | 23.2 | 23.3 | 29.0 | 24.9 |
| SC5778 | 30.2 | 30.6 | 30.1 | 34.8 | 31.9 |
| SC5779 | 33.3 | 32.8 | 32.0 | 36.5 | 34.7 |
| SC5780 | 14.6 | 13.7 | 13.9 | 19.2 | 15.1 |
| SC5781 | 15.1 | 14.1 | 14.3 | 20.2 | 16.2 |
| SC5782 | 21.8 | 20.9 | 21.0 | 26.9 | 22.6 |
| SC5783 | 16.0 | 14.9 | 15.6 | 20.8 | 16.9 |
| SC5784 | 36.0 | 35.6 | Negative | Negative | 35.4 |
| SC5785 | 27.8 | 27.3 | 27.4 | 32.7 | 28.9 |
| SC5786 | 23.9 | 24.0 | 24.3 | 29.4 | 25.6 |

### Assays using the CDC N2 and Corman E-gene primer/probe sets were more sensitive than those using the CDC N1 and Corman RDRP sets and the BGI kit

Our final analysis was to test the sensitivity and specificity of assays using the CDC primer/probe sets and compare these to the sensitivity and specificity of assays using the Corman RdRp and E-gene sets. Because assays using the N3 set did not perform as well as those using the N1 and N2 sets, we did not include the latter set in this analysis. We did, however, include in this analysis an evaluation of the sensitivity and specificity of assays performed using a SARS-CoV-2 test kit from BGI. To directly compare LODs between assays using the Corman RdRp and E-gene sets to those using the N1 and N2 sets and the BGI kit, we ran assays on dilutions ranging from 1:10^5^ to 1:10^7^ of the positive UW Virology clinical sample. We again ran twenty duplicate assays with each primer/probe set and with the BGI kit on each dilution. The results of the analysis are shown in Table 3. The least sensitive assays were the ones that used the Corman RdRp primer/probe set. At a dilution of 1:10^5^, only 17 out of the 20 assays that used this set were positive for SARS-CoV-2. The most sensitive assays used the CDC N2 and the Corman E-gene sets for which 18 and 17 replicate assays, respectively, were positive at a dilution of 1:10^6^.

**Table 3:** Relative sensitivities of SARS-CoV-2 detection assays using 4 different primer/probe sets and the BGI testing kit.

| Corman E-gene |  |  |  |  |  |  |  |
| --- | --- | --- | --- | --- | --- | --- | --- |
| Dilution | Sample ID | Replicates |  |  | % Positivity | CT<sup>1,2</sup><br>Mean | SD<sup>3</sup> |
|  |  | Tested | Positive | Negative |  |  |  |
| 1:10<sup>5</sup> | S5 | 20 | 20 | 0 | 100 | 33.7 | 1.13 |
| 1:10<sup>6</sup> | S6 | 20 | 17 | 3 | 85 | 37.2 | 1.34 |
| 1:10<sup>7</sup> | S7 | 20 | 3 | 17 | 15 | 37.9 | 0.10 |

| Corman RdRp gene |  |  |  |  |  |  |  |
| --- | --- | --- | --- | --- | --- | --- | --- |
| Dilution | Sample ID | Replicates |  |  | % Positivity | CT<br>Mean | SD |
|  |  | Tested | Positive | Negative |  |  |  |
| 1:10<sup>5</sup> | S5 | 20 | 17 | 3 | 85 | 36.6 | 1.84 |
| 1:10<sup>6</sup> | S6 | 20 | 2 | 18 | 10 | 35.8 | 1.79 |
| 1:10<sup>7</sup> | S7 | 20 | 1 | 19 | 5 | 37.5 | 0.00 |

| CDC N1 |  |  |  |  |  |  |  |
| --- | --- | --- | --- | --- | --- | --- | --- |
| Dilution | Sample ID | Replicates |  |  | % Positivity | CT<br>Mean | SD |
|  |  | Tested | Positive | Negative |  |  |  |
| 1:10<sup>5</sup> | S5 | 20 | 20 | 0 | 100 | 33.7 | 1.45 |
| 1:10<sup>6</sup> | S6 | 20 | 13 | 7 | 65 | 36.2 | 1.41 |
| 1:10<sup>7</sup> | S7 | 20 | 2 | 18 | 10 | 36.2 | 0.55 |

| CDC N2 |  |  |  |  |  |  |  |
| --- | --- | --- | --- | --- | --- | --- | --- |
| Dilution | Sample ID | Replicates |  |  | % Positivity | CT<br>Mean | SD |
|  |  | Tested | Positive | Negative |  |  |  |
| 1:10<sup>5</sup> | S5 | 20 | 20 | 0 | 100 | 33.1 | 1.26 |
| 1:10<sup>6</sup> | S6 | 20 | 18 | 2 | 90 | 36.8 | 1.35 |
| 1:10<sup>7</sup> | S7 | 20 | 3 | 17 | 15 | 37.2 | 0.61 |

| BGI Kit |  |  |  |  |  |  |  |
| --- | --- | --- | --- | --- | --- | --- | --- |
| Dilution | Sample ID | Replicates |  |  | % Positivity | CT<br>Mean | SD |
|  |  | Tested | Positive | Negative |  |  |  |
| 1:10<sup>5</sup> | S5 | 20 | 20 | 0 | 100 | 31.6 | 2.39 |
| 1:10<sup>6</sup> | S6 | 20 | 10 | 10 | 50 | 34.7 | 2.06 |
| 1:10<sup>7</sup> | S7 | 20 | 0 | 20 | 0 | - | - |

<sup>1</sup>Cycle-time
<sup>2</sup>Only positive results are included in calculation of mean CT
<sup>3</sup>Standard deviation

The specificities assays using the CDC N1 and N2 primer/probe sets and the BGI kit were also tested using our panel of samples positive for various respiratory viruses. Assays using the N1 and N2 sets and the BGI kit were found to be 100% specific.

## Conclusions

Known cases of COVID-19 have now exceeded 105,000 worldwide. While cases in China appear to be leveling off with fewer new cases diagnosed each day, new case clusters are rapidly appearing in other nations across the world (3). In the coming days and weeks, many clinical laboratories will be developing their own SARS-CoV-2 testing protocols. Maximizing the sensitivity and specificity of these tests is critical to efforts around the world to minimize the impact of this epidemic on global health.

A number of different primer/probe sets for use in SARS-CoV-2 detection assays and SARS-CoV-2 testing kits have been developed and are now available. As we have demonstrated here, the performance characteristics of assays using these primer/probe sets and testing kits are variable. Of the seven different primer/probe sets and one testing kit that we evaluated, all were found to be highly specific with no false positive results observed when assays were run on samples positive for a number of other respiratory viruses. Variability was, however, observed in the sensitivities of these tests. We found that assays using the CDC N2 and Corman E-gene primer/probe sets to be particularly sensitive. Assays using these sets were able to detect SARS-CoV-2 in ten out of ten known positive clinical samples. They were also able to reliably detect SARS-CoV-2 in a positive sample diluted down to 1:10^6^. In addition to our evaluation of different assays for SARS-CoV-2 detection, we also show that it is possible to significantly increase capacity for the RNA extraction step of SARS-CoV-2 testing without sacrificing sensitivity.

In summary, we report variable performance characteristics of assays using seven different primer/probe sets and one complete testing kit used for SARS-CoV-2 testing of clinical samples. While assays using all sets were highly specific, some, such as those using the CDC N2 and the Corman E-gene sets, were found to be more sensitive than others. These findings will provide important insights on SARS-CoV-2 detection assay design to labs that are currently working to develop their own testing methods. Our results also emphasize the importance of on-going optimization of viral detection assays following the emergence of novel viral pathogens.

## Data Availability

Data referred to in the manuscript has been included in the manuscript's tables.

